# Association of antihypertensive agents with the risk of in-hospital death in patients with Covid-19

**DOI:** 10.1101/2020.11.23.20237362

**Authors:** Laurent Chouchana, Nathanaël Beeker, Nicolas Garcelon, Bastien Rance, Nicolas Paris, Elisa Salamanca, Elisabeth Polard, Anita Burgun, Jean-Marc Treluyer, Antoine Neuraz, On behalf of AP-HP / Universities / Inserm COVID-19 research collaboration; AP-HP Covid CDR Initiative and ‘Entrepôt de Données de Santé’ AP-HP consortium

## Abstract

In this retrospective multicenter cohort study, we aimed to investigate the association between antihypertensive agent exposure and in-hospital mortality in patients with Covid-19. Of 8,078 hospitalized patients for Covid-19, 3,686 (45.6%) had hypertension including 2043 (55.4%) patients exposed to a renin-angiotensin-aldosterone inhibitors (RAASi), 1624 (44.1%) to calcium channel blockers (CCB) and 1154 (37.7%) to beta-blockers. Overall in-hospital 30-day mortality was 23.1%. Compared to non-users, the risk of mortality was lower in CCB (aOR, 0.83 [0.70-0.99]) and beta-blockers users (aOR, 0.80 [0.67-0.95]), and not different in RAASi users. These findings support the continuation of antihypertensive agents in patients with Covid-19.

## INTRODUCTION

The coronavirus disease-2019 (Covid-19), due to the novel severe acute respiratory syndrome coronavirus-2 (SARS-CoV-2) is a global pandemic representing a major treat to global health. Data from several cohorts have shown that age and cardiovascular comorbidities, including hypertension, were among the main determinants of severe disease.^1,2^ As SARS-CoV-2 uses the angiotensin-converting enzyme 2 (ACE2) receptor as an entrance door into cells, questions have been raised about the role of the renin-angiotensin-aldosterone system (RAAS) activity on Covid-19 pathophysiology.^3^ RAAS inhibitors directly acts on RAAS in two different ways, both leading to ACE2 upregulation.^3^ On the contrary, some authors have hypothesized that the use of RAAS inhibitors may protect against acute lung injury caused by angiotensin II accumulation.^4^ Therefore, since these drugs such as ACE inhibitors (ACEi) or angiotensin receptor blockers (ARB) are common treatments for hypertension, controversies about their role on Covid-19 severity have been raised. To date, scientific societies have advised against their discontinuation in patients with hypertension, until robust clinical evidence is available.^5^ Previous reports showed that RAAS inhibitors do not increase the risk of severe or fatal outcome in Covid-19 patients.^1,6,7^ The role of other antihypertensive agents is unclear. However, these analyses are retrospective and additional data are needed to ascertain the beneficial or harmful effect of antihypertensive agents on Covid-19 mortality. In order to provide additional information, we performed a multicenter retrospective cohort study in patients hospitalized for Covid-19 in Greater Paris area, France.

## METHODS

### Study population

This retrospective cohort study was conducted using the Assistance Publique-Hopitaux de Paris (AP-HP) Health Data Warehouse (‘Entrepôt de Données de Santé (EDS)’ https://eds.aphp.fr/). This data-warehouse contains electronic health records (EHRs) of all inpatients from the 39 greater Paris university hospitals. Adult patients having at least one SARS-CoV-2 positive PCR test and hospitalized between February 1^st^ and May 15^th^, 2020 have been included. Patients having a PCR test performed more than fourteen days after hospitalization have been excluded considering they were very likely to be nosocomial infection. Patients have been followed at least 30 days after inclusion (date of first PCR test positive). The Institutional Review Board of the AP-HP Health Data Warehouse approved this study on April 7^th^, 2020 (CSE-20-18_COVID19).

### Comorbidities and drug exposure

Hypertension as well as other comorbidities have been retrieved in EHRs using specific codes from the International Classification of Diseases 10^th^ version, combined with natural language processing (regular expressions) as previously described, within the six months previous to the hospital stay for Covid-19 (Suppl Table S1).^8^ Antihypertensive agent exposure have been identified in EHRs at admission of the hospital stay for Covid-19 and within the six previous months, using both specific Anatomical Therapeutic Chemical classification (ATC) codes and deep learning algorithm on clinical narratives as previously described (Suppl Table S2).^8^ As described in Neuraz et al.^8^, we extracted drug mentions and their attributes (dose, frequency, duration, route, condition) using a deep-learning pipeline based on fine-tuned multilingual embeddings and bidirectional long-short term memory units combined with conditional random fields.^9^ The drug mentions were then normalized to ATC using exact string matching.

### Statistical analysis

The primary outcome was all-cause 30-day in-hospital mortality. We assessed the association between antihypertensive agent class exposure and primary outcome using a multivariate logistic regression. Analyses were adjusted on age, sex and main chronic diseases to take into account confounding factors known to be associated with Covid-19 mortality. Data are presented as median (interquartile range [IQR]) and numbers (%). The results of the logistic regression model are presented as crude odds-ratio (OR) or adjusted OR (aOR) and their 95% confidence interval ([95% CI]). All analyses were performed using R statistical software.

## RESULTS

In Greater Paris University hospitals, 8,078 patients had been hospitalized for Covid-19 between February 1^st^ and May 15^th^ 2020, among which 1,531 (19.0%) died within 30 days. Among hospitalized patients, 3,686 (45.6%) had hypertension and were included in the study. In this population, median age was 75.4 (21.5) years and 57.1 % were male (Table 1). Regarding antihypertensive agents, the main pharmacological classes used were RAAS inhibitors (n=2043, 55.4%), calcium channel blockers (CCB) (n=1624, 44.1%), beta-blockers (n=1154, 37.7%), and centrally acting sympatholytics (n=172, 4.7%). More specifically for RAAS inhibitors, ARB and ACEi were used in 1154 (31.3%) and 998 (27.1%) patients, respectively, and no patient received a renin-inhibitor. In 826 patients (22.4%) no antihypertensive treatment was identified.

**Table 1.** Patient characteristics. These groups are not exclusive and one patient can be exposed to more than one pharmacological class. Non-survivors are considered in relation with in-hospital 30-day mortality. ARB: angiotensin II receptor blockers; ACEi: angiotensin-converting enzyme inhibitors; CCB: calcium channel blockers.

|  | **All patients** | **Patients with hypertension** |  |  |
| --- | --- | --- | --- | --- |
|  | **(N=8078)** | **All (N=3686)** | **Non survivors (N=853)** | **Survivors (N=2833)** |
| Age – years, median (IQR) | 68 (25.9) | 75.4 (21.5) | 82.1 (16.3) | 72.9 (22.2) |
| Age – distribution |  |  |  |  |
| 18-44 years | 1020 (12.6%) | 123 (3.3%) | 5 (0.6%) | 118 (4.2%) |
| 45-64 | 2544 (31.5%) | 876 (23.8%) | 83 (9.7%) | 793 (28%) |
| 65-74 | 1662 (20.6%) | 813 (22.1%) | 165 (19.3%) | 648 (22.9%) |
| 75-84 | 1457 (18%) | 919 (24.9%) | 264 (30.9%) | 655 (23.1%) |
| > 85 | 1395 (17.3%) | 955 (25.9%) | 336 (39.4%) | 619 (21.8%) |
| Sex |  |  |  |  |
| Female | 3291 (40.7%) | 1583 (42.9%) | 330 (38.7%) | 1253 (44.2%) |
| Male | 4787 (59.3%) | 2103 (57.1%) | 523 (61.3%) | 1580 (55.8%) |
| Chronic diseases |  |  |  |  |
| Hypertension | 3686 (45.6%) | 3686 (100%) | 853 (100%) | 2833 (100%) |
| Chronic kidney disease | 1644 (20.4%) | 1407 (38.2%) | 384 (45%) | 1023 (36.1%) |
| Cerebrovascular disease | 1511 (18.7%) | 1209 (32.8%) | 309 (36.2%) | 900 (31.8%) |
| Cardiovascular disease | 1762 (21.8%) | 1438 (39%) | 378 (44.3%) | 1060 (37.4%) |
| Cardiac failure | 1099 (13.6%) | 949 (25.7%) | 261 (30.6%) | 688 (24.3%) |
| Diabetes | 2220 (27.5%) | 1730 (46.9%) | 404 (47.4%) | 1326 (46.8%) |
| Respiratory disease | 1427 (17.7%) | 944 (25.6%) | 162 (19.0%) | 782 (27.6%) |
| Obesity | 1851 (22.9%) | 1072 (29.1%) | 239 (28.0%) | 833 (29.4%) |
| Malignancies | 1803 (22.3%) | 1286 (34.9%) | 324 (38.0%) | 962 (34.0%) |
| Antihypertensive drugs |  |  |  |  |
| RAAS inhibitors | 2720 (33.7%) | 2043 (55.4%) | 441 (51.7%) | 1602 (56.5%) |
| ARB | 1520 (18.8%) | 1154 (31.3%) | 250 (29.3%) | 904 (31.9%) |
| ACEi | 1337 (16.6%) | 998 (27.1%) | 214 (25.1%) | 784 (27.7%) |
| CCB | 2210 (27.4%) | 1624 (44.1%) | 340 (39.9%) | 1284 (45.3%) |
| Beta-blockers | 1920 (23.8%) | 1389 (37.7%) | 315 (36.9%) | 1074 (37.9%) |
| Centrally acting sympatholytics | 259 (3.2%) | 172 (4.7%) | 35 (4.1%) | 137 (4.8%) |
| No antihypertensive drug | 3982 (49.3%) | 826 (22.4%) | 229 (26.8%) | 597 (21.1%) |

In total, there were 853 (23.1%) all-cause in-hospital 30-day deaths. Patients who died were older and there were more likely to be men, compared to survivors. They also had chronic diseases more frequently, except for respiratory diseases and obesity. The risk of mortality was lower in CCB (aOR, 0.83 [0.70-0.99]) and beta-blockers users (aOR, 0.80 [0.67-0.95]), and non-significant in ARB (aOR, 0.88 [0.72-1.06]) and ACEi users (aOR, 0.83 [0.68-1.02]), compared to non-users (Figure 1, Suppl. Table S3).

**Figure 1.**
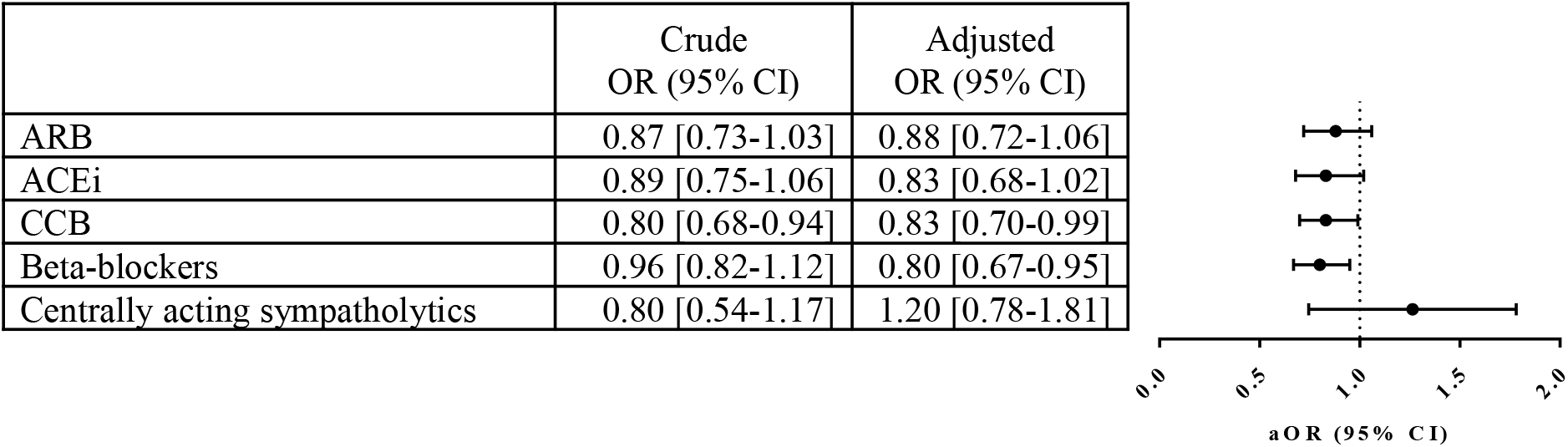
In-hospital 30-day mortality in hypertensive patients with Covid-19 according to drug exposure. These groups are not exclusive and one patient can be exposed to more than one pharmacological class. Analyses have been adjusted on age, sex and main chronic diseases (i.e. hypertension, chronic kidney disease, cerebrovascular disease, cardiovascular disease, cardiac failure, diabetes, respiratory disease, obesity and malignancies). ARB: angiotensin II receptor blockers; ACEi: angiotensin-converting enzyme inhibitors; CCB: calcium channel blockers

## DISCUSSION

This study, one of the largest multicenter retrospective to date on more than 3600 hospitalized Covid-19 patients with hypertension, provides two main findings. First, based on more than 2,000 patients exposed to RAAS inhibitors, there is no association with the use of ACEi or ARB and the risk of in-hospital death. Second, there is a significant protective effect of calcium-channel blockers (CCB) or beta-blocker use on the risk of death in hypertensive patients with Covid-19.

Hypertension and cardiovascular comorbidities have been previously reported as risk factors for severe disease and fatal outcome.^1,10,11^ The underlying pathogenic mechanisms of these comorbidities remain unclear and may involve the RAAS as being a double-edged sword. On one hand, RAAS inhibitors increase the expression of ACE2, which could promote virus entry.^3^ On the other hand, RAAS inhibitors, especially ARB, could reverse deleterious effects of unopposed angiotensin II accumulation resulting from ACE2 down-regulation associated to SARS-CoV-2 entry in cells.^4,12^ Clinical evidence based on a large study including 12,594 tested patients for SARS-CoV-2 showed no increase of likelihood to have a positive test among RAAS users.^13^ Furthermore, in line with our findings, a meta-analysis on four retrospective studies that include 921 ACEi or ARB users found no difference in risk of death compared to non-users (OR, 0.88 [0.68-1.14]).^14^

In our study, we found that CCB and beta-blockers were more frequently used in survivors (OR 0.83 [0.70-0.99] and 0.80 [0.67-0.95], respectively) than in non-survivors. These findings were statistically significant. Previous Covid-19 cohort studies did not found any beneficial or deleterious effect of CCB use on Covid-19 severity or mortality.^7,15^ However, these studies included few CCB users and were probably lacking power. Recently, an *in vitro* study on an emerging porcine coronavirus showed that CCB could prevent from infection and intracellular calcium homeostasis dysregulation.^16^ Regarding beta-blockers, one could hypothesizes that they can counteract deleterious sympathetic activation during the SARS-CoV-2 induced cytokine storm and severe disease. Further clinical data are needed to explore these early findings.

Our study has some limitations. First, in this retrospective observational design on hospitalized Covid-19 patients, hypertension or antihypertensive exposure could have affected the chance of hospitalization, which may limit the generalizability of our results. Second, although analyses were adjusted on major comorbidities, some unmeasured or unknown confounders may have not been ruled out, including indication bias. Finally, ascertainment of medication use from EHRs may not capture all drug prescriptions and not reflect actual drug exposure. However, considering antihypertensive agents are chronic treatments, we consider exposure to these drugs up to six months prior to hospitalization for Covid-19.

## CONCLUSION

In summary, we did not find any association between the use of RAAS inhibitors and the risk of in-hospital death in Covid-19 patients with hypertension. Interestingly we found a reduced mortality among CCB and beta-blockers users. Further studies are needed to confirm this protective role. Our findings, such as previous reports, support the continuation of antihypertensive agents in Covid-19 patients, in line with the current guidelines.

## Data Availability

Data available on request. The data underlying this article will be shared on reasonable request to the corresponding author.

## DECLARATIONS

## Acknowledgments

The authors thank the EDS AP-HP Covid consortium integrating the AP-HP Health Data Warehouse team as well as all the AP-HP staff and volunteers who contributed to the implementation of the EDS-Covid database and operating solutions for this database.

## Author contribution statement

Study design: LC, AN

Data management and database build process: NG, BR, NP, ES

Statistical analysis: NB, AN

Result interpretation: LC, AN

Draft the manuscript: LC

Critically review the manuscript: LC, AN, EP, AB, JMT

Approved the manuscript: LC, NB, NG, BR, NP, ES, EP, AB, JMT, AN

## Collaborators

**AP-HP/Universities/INSERM COVID-19 Research Collaboration; AP-HP COVID CDR Initiative** Collaborators: Pierre-Yves ANCEL, Alain BAUCHET, Nathanaël BEEKER, Vincent BENOIT, Mélodie BERNAUX, Ali BELLAMINE, Romain BEY, Aurélie BOURMAUD, Stéphane BREANT, Anita BURGUN, Fabrice CARRAT, Charlotte CAUCHETEUX, Julien CHAMP, Sylvie CORMONT, Christel DANIEL, Julien DUBIEL, Catherine DUCLOAS, Loic ESTEVE, Marie FRANK, Nicolas GARCELON, Alexandre GRAMFORT, Nicolas GRIFFON, Olivier GRISEL, Martin GUILBAUD, Claire HASSEN-KHODJA, François HEMERY, Martin HILKA, Anne Sophie JANNOT, Jerome LAMBERT, Richard LAYESE, Judith LEBLANC, Léo LEBOUTER, Guillaume LEMAITRE, Damien LEPROVOST, Ivan LERNER, Kankoe LEVI SALLAH, Aurélien MAIRE, Marie-France MAMZER, Patricia MARTEL, Arthur MENSCH, Thomas MOREAU, Antoine NEURAZ, Nina ORLOVA, Nicolas PARIS, Bastien RANCE, Hélène RAVERA, Antoine ROZES, Elisa SALAMANCA, Arnaud SANDRIN, Patricia SERRE, Xavier TANNIER, Jean-Marc TRELUYER, Damien VAN GYSEL, Gaël VAROQUAUX, Jill Jen VIE, Maxime WACK, Perceval WAJSBURT, Demian WASSERMANN, Eric ZAPLETAL.

