## Supplemental data for "Association of antihypertensive agents with the risk of in-hospital death in patients with Covid-19"

**Table S1. International classification of disease (10^th^ edition) codes and regular expressions used for comorbidities**

|  | ICD-10 codes | Regexp |
| --- | --- | --- |
| Hypertension | I10-I13, I15 | "[^A-Z]HTA[^A-Z]\|hyperten[st]ion.{0,5}art[eé]rielle\|hyperten[st]ion.{0,5}[eé]s{1,2}entiel{1,2}." |
| Chronic kidney disease | N18 | "maladie.{0,5}r[eé]nale\|n[eé]phropathie\|Insuffisance.{0,5}r[eé]nale.{0,5}chronique\|[^a-z]IRC[^a-z]\|[^a-z]dialyse\|h[ée]modialyse" |
| Cerebrovascular disease | I60-I67, I69 | "[^a-z]AVC[^a-z]\|acc*ident.{0,5}vasculaire.{0,5}c[ée]r[ée]bral\|acc*ident.{0,5}isch?[ée]mique.{0,5}c[ée]r[ée]bral\|acc*ident.{0,3}h[ée]morragique.{0,3}c[ée]r[ée]bral" |
| Cardiovascular disease | I20-I25 | "[Aa]tteinte.{0,5}cardiaque\|[Vv]alvule\|[Cc]ardiaque\|[Cc]ardiopathie\|[Nn].phropathie\|[Hh]ypertens\|[Ee]mbolie\|cardiopulmo\|cardite\|vasomotrice" |
| Cardiac failure | I50 | "insuf{1,}isance.{0,5}cardiaque\|d[ée]faill[ea]nce.{0,5}cardiaque" |
| Diabetes | E10-E14 | "(?<!pre)(?<!pre.)diab[ée]te\|[^a-z]DID[^a-z]\|[^a-z]DNID[^a-z]\|diab[ée]tique" |
| Respiratory disease | J44, J45 | "a[sth][sth][sth]me\|bronch?ospasme\|a[sth][sth][sth]matique",  "insuf.{1,2}isance.{0,5}respiratoire.{0,5}chronique",  "emph.s[èe]me\|emf.s[èe]me" |
| Obesity | E66 | "ob[éeè]sit[éeè]\|ob[èeé]se" |
| Malignancies | C00-C97, D00-D48 | "(?<!pre)(?<!pre )cancer[^a-z]\|tumeur(?!.{0,7}benigne)\|carcinome\|m[eé]lanome\|n[eé]oplasie\|sarcome"  "Lymphome"  "Leuc[ée]mie"  "leuc[ée]mie\|lymphome\|my[ée]lome" |

**Table S2. Anatomical Therapeutic Chemical classification codes used for drug exposure**

|  | **Plain or in combination with diuretics, including thiazides)** | **Fixed combination with other drugs** |
| --- | --- | --- |
| **ACEi** | C09A, C09BA, C09BX01, C09BX03 | C09BB, C09BX02, C09BX04 |
| **ARB** | C09C, C09DA, C09DX01, C09DX03, C09DX06, C09DX07 | C09DB, C09DX05 |
| **Centrally acting sympatholytics** | C02A, C02LA, C02LB, C02LC | - |
| **CCB** | C08C, C08GA, C09BX01, C09BX03, C09DX01, C09DX03, C09DX06, C09DX07 | C09BB, C09BX04, C09DB, C07FB |
| **Beta-blockers** | C07A, C07FX, C07B, C07C, C07D | C09BX02, C09BX04, C09DX05, C07FB |

ACEi: angiotensin-converting enzyme inhibitors; ARB: angiotensin II receptor blockers; CCB: calcium channel blockers

**Table S3. Association between all-cause 30-day mortality and age, sex and major comorbidities**

|  | **Crude OR (95% CI)** | **Adjusted OR (95% CI)** |
| --- | --- | --- |
| Age – year |  |  |
| 18-44 | Reference | Reference |
| 45-64 | 2.37 [1.04-6.85] | 2.35 [1.02-6.82] |
| 65-74 | 5.87 [2.61-16.8] | 6.11 [2.70-17.56] |
| 75-84 | 9.26 [4.15-26.4] | 10.43 [4.62-29.91] |
| > 85 | 12.4 [5.56-35.32] | 15.97 [7.06-45.88] |
| Sex |  |  |
| Female | Reference | Reference |
| Male | 1.26 [1.07-1.47] | 1.71 [1.44-2.04] |
| Chronic diseases |  |  |
| Chronic kidney disease | 1.45 [1.24-1.69] | 1.28 [1.07-1.53] |
| Cerebrovascular disease | 1.21 [1.03-1.43] | 1.03 [0.86-1.22] |
| Cardiovascular disease | 1.32 [1.13-1.54] | 1.11 [0.93-1.33] |
| Cardiac failure | 1.36 [1.14-1.61] | 1.00 [0.82-1.22] |
| Diabetes | 1.02 [0.87-1.19] | 1.08 [0.91-1.29] |
| Respiratory disease | 0.61 [0.50-0.73] | 0.54 [0.44-0.66] |
| Obesity | 0.95 [0.80-1.13] | 1.46 [1.20-1.77] |
| Malignancies | 1.21 [1.03-1.42] | 1.03 [0.87-1.22] |
